# Comparative IgG responses to SARS-CoV-2 after natural infection or vaccination

**DOI:** 10.1101/2022.09.28.22280476

**Authors:** Kaylan M. Olds, Devon P. Humphreys, Kathleen M. Gavin, Anne L. Wyllie, Timothy A. Bauer

**Author notes:** Corresponding Author **Address for correspondence:** Timothy Bauer, PhD, Everly Health, Inc., 823 Congress Ave., Suite 1200, Austin, TX 78701.

## Abstract

**Background:** Whether vaccination or natural infection provides greater benefit regarding the development of sustained immunity against SARS-CoV-2 remains unknown. Therefore, the aim of this study was to provide a direct comparison of IgG durability in vaccinated and unvaccinated adults.

**Methods:** This was a prospective, cross-sectional study of antibody durability in 1087 individuals with a median (IQR) age of 42 (35, 52) years who were unvaccinated and previously infected with SARS-CoV-2 (Arm 1, n=351) or vaccinated against the virus (Arm 2, n=737). Participants self-reported vaccination and infection history and provided self-collected serology samples using mailed collection kits.

**Results:** Anti-S1 IgG seroprevalence was 15.6% higher in vaccinated versus unvaccinated, previously-infected individuals across intervals ranging from 1 to 12 months and antibody survival was sustained near 100% through 12 months in the vaccinated group.

**Conclusions:** These findings suggest that vaccination as opposed to natural infection alone provides significant advantages in terms of sustained and effective immunity against prior variants of SARS-CoV-2. Future efforts to characterize SARS-CoV-2 immune responses should address hybrid immunity, booster status and formulation, and protection against (sub)variants of Omicron and future lineages, as well as weigh the potential impact of other immune system mechanisms.

## Introduction

Throughout the course of the Coronavirus Disease 2019 (COVID-19) pandemic, the widespread adoption of vaccination has been promoted for its potential to significantly curtail health and economic burdens related to the spread of SARS-CoV-2, and ultimately, key to bringing the pandemic to an end (1, 2). In less than a year from the World Health Organization (WHO) declaring the pandemic, following an unprecedented development effort, vaccines were available to most adults in western societies. However, vaccination campaigns were met with varying degrees of hesitancy in portions of the population (3-5). In the intervening months, researchers documented Immunoglobulin G (IgG) seroconversion and fluctuating levels of sustained immunity resulting from natural infection and/or vaccination (6-15), and established a positive relationship between anti-spike antibodies and clinical protection from SARS-CoV-2 (16-18).

However, the current literature does not provide a clear difference in antibody profiles acquired through vaccination and natural infection. This distinction is of renewed importance at a time when nearly 60% of the United States (US) population (including 75% of US children) are reported to show serological evidence of community exposure (19) and the sense of urgency related to vaccination has waned. Despite a growing body of literature, availability of data directly comparing antibody responses following vaccination versus natural infection is limited. Prospective evaluations tend to be limited in sample size (12, 14, 20-22), have short serological monitoring periods,(9, 20, 22) or analyses were completed retrospectively (23, 24). Many studies utilize populations of healthcare workers who may encounter infectious agents – including SARS-CoV-2 – more frequently than other members of the community, limiting generalizability of results due to potential repeated exposures and subclinical infections (9, 12, 20, 25). Several quality studies have evaluated the effectiveness of vaccination- and/or infection-acquired immunity over time, but have lacked a serological component (24-26). Therefore, in the current study, we aimed to provide a direct comparison of IgG durability in vaccinated and unvaccinated individuals in a prospective US sample. This work presents the largest such comparison in a prospectively-collected, population-wide sample to date.

## Methods

### Ethics approval and consent to participate

This protocol was approved by WCG IRB (IRB registration #20210763). All participants provided written informed consent prior to enrollment in the study.

### Study design

This was a prospective, cross-sectional study of antibody durability in individuals who were previously infected with SARS-CoV-2 (Arm 1) or vaccinated against the virus (Arm 2). The study utilized electronic questionnaires and self-collected dried blood spot (DBS) samples to gather data remotely from participants within the US. Upon enrollment, participants answered questions about COVID-19 diagnosis and vaccination history, symptom and treatment information, and other relevant medical history such as comorbid conditions and medication use. Participants received a COVID-19 Antibody Home Collection Kit (Everlywell, Inc., Austin, TX) to provide samples for qualitative IgG detection. Baseline data were captured to complete the primary analysis of comparative IgG durability between study arms. In addition, Arm 2 participants repeated questionnaires and serology tests during a follow-up period for up to 9 months, facilitating a secondary survival analysis of vaccine-acquired IgG.

### Participant eligibility and enrollment

Participants were enrolled in the study between March and November 2021. Inclusion criteria for Arm 1 included prior infection with SARS-CoV-2 (evidenced by a positive diagnostic test) and unvaccinated status. Eligibility for Arm 2 required receipt of at least one dose of a SARS-CoV-2 vaccine. All participants were required to be age 18 years or older, reside within the continental US, and have access to an email account and internet service. Exclusion criteria included known conditions or ongoing treatments associated with immune impairment (e.g., chemotherapy) and residents of New York state. All participants provided written informed consent prior to enrollment in the study, which was approved by WCG IRB (IRB study #20210763).

### Serology Testing

DBS samples were self-collected at baseline and follow-up timepoints for serology testing using the Anti-SARS-CoV-2 ELISA Assay (EUROIMMUN, Germany).

### Study Endpoints

The primary endpoint was the expected difference in S1-binding IgG seropositivity between Arm 1 (unvaccinated, naturally infected individuals) and Arm 2 (vaccinated individuals) over time. The secondary endpoints included: S1-binding IgG survival percentages at 4 and 12 months since last exposure to a vaccine product in Arm 2.

### Statistical Analysis

Multiple linear regression was used to evaluate the effect size and significance of each study arm (vaccination vs natural infection) on estimated seropositivity over time (binned as the number of months since last vaccine dose or infection). Estimates per study arm per time interval that included fewer than 3 serology values were considered under sampled and dropped prior to analysis.

Additionally, a discrete-time analysis was conducted using independent z-tests of proportions at 1, 3, 6, 9, and 12 months to evaluate differences in seropositivity between Arms 1 and 2. P-values are reported and interpretable using a Bonferroni-adjusted value of 0.005. To further evaluate S1-binding IgG durability associated with vaccination, survival analysis was performed using the Kaplan-Meier method. Percent survival at 4 and 12 months is reported with 95% confidence intervals. This analysis was conducted in two ways, reflecting (1) time from the most recent vaccination dose (i.e., a booster dose reset the interval clock), and (2) time from the initial vaccination dose.

## Results

### S1-binding IgG Cross-Sectional Analysis

A total of 1,087 participants enrolled in the study and completed all required baseline assessments for inclusion in the analysis: 351 were assigned to Arm 1 and 737 were assigned to Arm 2 at baseline. Over three quarters of participants included in the cross-sectional analysis were female, and the median (IQR) age was 42 (35, 52) years. Over two thirds of vaccinated participants reported no previous SARS-CoV-2 infection prior to entering the study. Baseline participant characteristics are summarized in Table 1.

**Table 1:** Participant demographics and diagnostic test history

| Characteristic | Overall<br>N = 1088 | Arm 1<br>N = 351 | Arm 2<br>N = 737 |
| --- | --- | --- | --- |
| **Sex** |  |  |  |
| Female | 831 (76%) | 258 (74%) | 573 (78%) |
| Male | 255 (23%) | 93 (26%) | 162 (22%) |
| Unknown | 2 (0.2%) | 0 (0%) | 2 (0.3%) |
| **Age** |  |  |  |
| Median (IQR) | 42 (35, 52) | 43 (34, 54) | 42 (35, 51) |
| Unknown | 2 (0.2%) | 0 (0%) | 2 (0.3%) |
| **COVID Diagnostic History** |  |  |  |
| Known Infection | 566 (52%) | 351 (100%) | 215 (29%) |
| No Known Infection | 508 (47%) | 0 (0%) | 508 (69%) |
| Unknown/NA | 14 (1.3%) | 0 (0%) | 14 (1.9%) |
| **Vaccine Product** |  |  |  |
| Ad26.COV2.s (Johnson & Johnson) | 56 (5.1%) | 0 (0%) | 56 (7.6%) |
| BNT162b2 (Pfizer) | 396 (36%) | 0 (0%) | 396 (54%) |
| mRNA-1273 (Moderna) | 280 (26%) | 0 (0%) | 280 (38%) |
| NVX-CoV2373 (Novavax) | 1 (<0.1%) | 0 (0%) | 1 (0.1%) |
| Not Applicable | 351 (32%) | 351 (100%) | 0 (0%) |
| Unknown | 4 (0.4%) | 0 (0%) | 4 (0.5%) |

Nearly all Arm 2 participants entered the study fully vaccinated, defined as having received all doses in the primary series of the applicable vaccine, with only 3.7% reporting partial vaccination (or only having received one dose of a two-dose primary vaccination regimen) at baseline. No participants reported receiving a booster dose prior to enrollment. The difference in time between participants’ most recent confirmed exposure (positive diagnostic test date or vaccination date for Arm 1 or 2, respectively) and baseline serology test date ranged from 1 to 15 months (median) after dropping under-sampled time intervals (N=1,007).

Population-wide seropositivity remained high across time intervals. The seropositivity effect size associated with vaccination was 15.6% greater than the reference arm (Arm 1), independent of time since vaccination (Figure 1 and Table 2).

**Figure 1.**
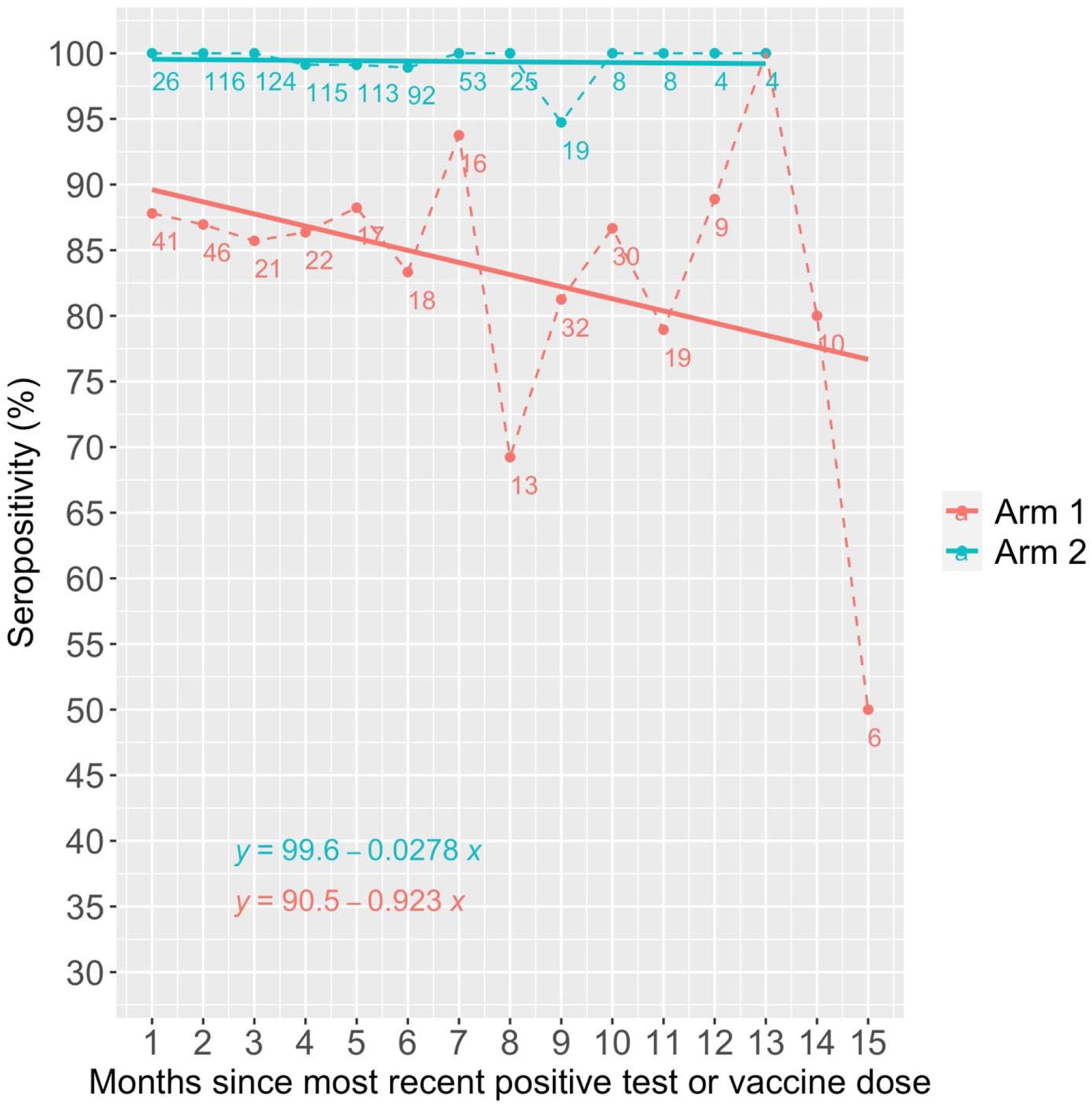
Anti-S1 seropositivity in unvaccinated/naturally infected (Arm 1) and vaccinated (Arm 2) groups

**Table 2:** Difference in seropositivity by exposure mode

| Time Interval | Difference in Arm 2 (Vaccinated) versus Arm 1 (Unvaccinated/Natural Infection) | Z-score | p-value |
| --- | --- | --- | --- |
| 1 month | +11.9% | -28.06 | $1.37 \times 10^{-173}$ |
| 3 months | +14.3% | -125.62 | 0.00 |
| 6 months | +15.6% | -66.93 | 0.00 |
| 9 months | +13.5% | -13.58 | $2.71 \times 10^{-42}$ |
| 12 months | +11.1% | -4.33 | $7.34 \times 10^{-6}$ |

### S1-binding IgG Longitudinal Analysis in vaccinated individuals (Arm 2)

A total of 1,605 samples from 737 vaccinated individuals were available for antibody survival analysis. Antibody survival (with 95% CI) ranged from 99.4% (99.0%-100%) at 4 months to 95.5% (91.7%-99.5%) at 12 months when measured in time since the last dose received, and from 99.7% (99.3% - 100%) at 4 months to 98.4% (97.1%-99.7%) at 12 months when measured from the first dose. No further loss of IgG detectability was observed beyond 12 months in either analysis.

## Discussion

This study aimed to provide the first direct comparison of IgG durability in vaccinated and unvaccinated individuals in a large, prospective, population-wide US sample. Both vaccination against COVID-19 and natural infection with SARS-CoV-2 are associated with seroconversion in most people. However, we found IgG seropositivity was significantly higher, more consistent, and declined less rapidly in vaccinated individuals than in those who were unvaccinated and previously infected with the virus. This pattern was evident one-month post-exposure (the earliest time interval evaluated) and persisted through 12 months post infection or most recent vaccination dose. One possible explanation for the discrepancy between vaccinated and naturally infected groups could be the controlled dose of target antigen provided (indirectly, in many cases) through vaccination, versus the inconsistent “dose” of various viral antigens acquired through natural infection. In a within-subject survival analysis, vaccine-acquired IgG antibodies persisted for up to 12 months (or more) in nearly all cases. This provides within-subject support for strong antibody durability out to 12 months when compared to the vaccinated group (Arm 2) in the cross-sectional analysis. Collectively, these analyses suggest that IgG antibodies raised through vaccination are more reliably durable than those induced by a natural SARS-CoV-2 infection in most individuals.

Compared to prior research, our estimated rate of seropositivity decline in the unvaccinated group was steeper. Alfego et al. found seropositivity decline to be approximately -0.004% per day, or approximately -0.12% per month (11), while we observed a decline of -0.93% per month. This may be reflective of our smaller sample size or due to an important distinction between each studies’ methods. Alfego et al. evaluated seropositivity from the time of a participant’s first recorded infection (positive diagnostic test) and did not take subsequent exposure events into account, whereas in the current study, we measured seropositivity from the time of the most recent confirmed infection. Nevertheless, the rate of seropositivity decline in the vaccinated cohort was much lower than in the unvaccinated cohort. This holds true whether we compare this rate to those estimates from Alfego et al. or to our own rate estimates for Arm 1 in the present study. Collectively, this suggests that S-protein antibodies raised through vaccination are more reliably durable than those induced by a natural SARS-CoV-2 infection in most individuals.

While neutralizing antibody (NAb) titer is a closer surrogate for immune protection, Anti-S IgG testing offers an advantage in scalability as the requisite immunoassays are less burdensome in terms of time, cost, complexity, and availability than functional NAb assays. Published works by Harvala et al. and Lumley et al. lend support to the clinical relevance of IgG values. The former reported that detection of IgG with the Euroimmun assay highly correlated with NAb titers above 1:100 in convalescent blood samples (27), while the latter observed a connection between S-binding IgG and protection from reinfection with earlier variants of SARS-CoV-2 in previously-infected healthcare professionals followed up to 6 months (18).

Given the correlation of IgG levels with NAb titers and that NAb concentration correlates with protection against infection and severe disease (16, 17), our observations suggest that vaccine-acquired immunity may also be more protective than an immune response triggered by natural infection alone. Our findings support similar observations made in Congolese individuals at 2 months post vaccination (with Ad26.COV.2 or BBIP-CorV) compared to 2, 3, or 6 months post natural infection (21). However, whether a vaccine-induced IgG response provides an advantage over the longer term remains unclear. For example, in a previous evaluation of purely vaccine-induced NAb activity in 62 healthcare workers, which excluded cases of prior exposures or breakthrough infections, Decru et al. found significant waning of neutralization activity 10 months after receiving a second dose of BNT162b2 (12). If NAb follows similar patterns as anti-S IgG, it may be that hybrid immunity provides the most durable NAb response. More research is needed to clarify which approach produces the longest enduring neutralization capability. Furthermore, the limitation remains that without more prevailing use of a common reference standard it is difficult to determine what level of titer is necessary for protection, whether that involves full immunity or simply protection from severe disease. In addition, the contribution of other immune factors (e.g., IgA and mucosal immunity) on overall protection should not be overlooked, especially considering those responses may follow different patterns than IgG after vaccination or natural infection.

One problem that warrants further study is the impact of symptom severity on antibody responses in previously-infected individuals who become vaccinated (hybrid immunity) compared to those who remain unvaccinated, given that binding IgG and NAb responses during and after severe disease are higher functioning and longer lasting than when symptoms are mild (28-30). Future research may also look at the relationships between viral variants and population-wide antibody profiles in vaccinated and unvaccinated cohorts, as well as how antibody characteristics connect to clinical outcomes and transmissibility in each group. Moreover, additional focus should be paid to hybrid immunity and the impact of repeated (or breakthrough) infections and boosters, including extended follow-up (8, 20) for antibody durability and titer beyond one year from each type of exposure in the general population. Flexible study designs that can more easily accommodate adjustments made in response to a rapidly changing viral variant landscape would help future studies maintain relevance over time as SARS-CoV-2 transitions from pandemic to endemic circulation.

### Limitations

The data for Arm 1 are cross-sectional, therefore individual rates of seroreversion could not be evaluated in direct comparison to Arm 2. Additionally, limited sample sizes at later time intervals reduced statistical confidence in some time-specific point estimates of seropositivity, and limited cases with confirmed hybrid immunity prevented us from evaluating those individuals as a separate group. Furthermore, this study relied on self-reported infection and vaccination history, and only took confirmed exposures (those that resulted in a positive diagnostic test) into account. There is a high likelihood that additional exposures to SARS-CoV-2 went undetected and were not reflected in the self-reported COVID-19 diagnostic data; however, we expect that such events would have inflated serology values most frequently in the unvaccinated group. As such, these findings give a conservative estimate of the differences between vaccination and natural infection in terms of antibody response. Finally, our study largely coincided with circulation of the Delta variant, and reflected immune responses triggered by exposure to Alpha, Beta, or Delta variants in previously-infected individuals. The Omicron (BA.1) variant was discovered as this study concluded and has exhibited greater transmissibility than previous variants due to a highly mutated spike protein and consequent immune evasion and increased receptor affinity (31). Subsequent sublineages (BA.1.1, BA.2, BA.3, BA.4, and BA.5) have since been identified and show similar or greater transmissibility and evasion of antibodies compared to the parent variant (32, 33). Furthermore, a new vaccine formulation that offers more protection against Omicron has been made available as a booster dose. It is uncertain how combinations of variant/subvariant exposures and primary/booster vaccine formulations may affect immune responses acquired through natural infection, vaccination, or hybrid exposure in a rapidly shifting landscape that continues to challenge the pace of science.

## Conclusions

This study was one of the first to directly compare population-level antibody durability acquired through vaccination or natural infection in a nationwide sample and supports the conclusion that completing the primary series of vaccination triggers a more durable and protective antibody response compared to a known single infection with early variants of SARS-CoV-2. This study further supports the importance of broad vaccination campaigns over relying upon natural, infection-driven herd immunity alone in the fight against SARS-CoV-2. Future work investigating antibody profiles resultant to booster doses, hybrid immunity, as well as repeat and variant specific infections are critical in our continued understanding of SARS-CoV-2 and COVID-19.

## Data Availability

The datasets used and/or analyzed during the current study are available from the corresponding author on reasonable request.

## Abbreviations

COVID-19: Coronavirus Disease 2019
DBS: dried blood spot
IgG: Immunoglobulin G
IQR: Interquartile range
NAb: Neutralizing antibody
US: United States
WHO: World Health Organization

## Declarations

### Consent for publication

Not applicable.

### Competing interests

Kaylan Olds, Devon Humphreys, Kathleen Gavin, and Timothy Bauer are full time employees of Everly Health, Inc. Anne Wylie has no conflicts to report.

### Funding

This study was funded and conducted by Everly Health, Inc.

### Authors’ contributions

KMO: Had a role in study conception, data interpretation, and drafted the work, DPH: Had a role in study conception, performed data analysis and interpretation, and assisted in drafting the work, KMG: Performed data interpretation and assisted in drafting the work, ALW: Had a role in data analysis, data interpretation, and assisted in drafting the work, TAB: Led study conception, data interpretation, and assisted in drafting the work. All authors have reviewed and approved this submitted version of the manuscript.

## Acknowledgements

The authors thank PerkinElmer Genomics, Inc. for providing in-kind support with serology testing.

## Notes

### Summary of Updates

Due to data sharing agreements with our sub-study laboratory partner, a portion of the manuscript has been retracted while the vendor reviews the material. This revision does not impact the primary analysis or conclusions of the manuscript, and we have removed the substudy portion from the current draft to avoid confusion.

## References

1. Iboi EA, Ngonghala CN, Gumel AB. Will an imperfect vaccine curtail the COVID-19 pandemic in the U.S.? Infect Dis Model. 2020;5:510–24.

2. Shah SMA, Rasheed T, Rizwan K, Bilal M, Iqbal HMN, Rasool N, et al. Risk management strategies and therapeutic modalities to tackle COVID-19/SARS-CoV-2. J Infect Public Health. 2021;14(3):331–46.

3. Kreps S, Prasad S, Brownstein JS, Hswen Y, Garibaldi BT, Zhang B, et al. Factors Associated With US Adults’ Likelihood of Accepting COVID-19 Vaccination. JAMA Netw Open. 2020;3(10):e2025594.

4. Ball P. Anti-vaccine movement could undermine efforts to end coronavirus pandemic, researchers warn. Nature. 2020;581(7808):251.

5. Szilagyi PG, Thomas K, Shah MD, Vizueta N, Cui Y, Vangala S, et al. National Trends in the US Public’s Likelihood of Getting a COVID-19 Vaccine-April 1 to December 8, 2020. JAMA. 2020.

6. Goldberg Y, Mandel M, Bar-On YM, Bodenheimer O, Freedman LS, Ash N, et al. Protection and Waning of Natural and Hybrid Immunity to SARS-CoV-2. N Engl J Med. 2022;386(23):2201–12.

7. Amirthalingam G, Bernal JL, Andrews NJ, Whitaker H, Gower C, Stowe J, et al. Serological responses and vaccine effectiveness for extended COVID-19 vaccine schedules in England. Nat Commun. 2021;12(1):7217.

8. Prendecki M, Clarke C, Brown J, Cox A, Gleeson S, Guckian M, et al. Effect of previous SARS-CoV-2 infection on humoral and T-cell responses to single-dose BNT162b2 vaccine. Lancet. 2021;397(10280):1178–81.

9. Payne RP, Longet S, Austin JA, Skelly DT, Dejnirattisai W, Adele S, et al. Immunogenicity of standard and extended dosing intervals of BNT162b2 mRNA vaccine. Cell. 2021;184(23):5699-714.e11.

10. McDade TW, Sancilio A, D’Aquila R, Mustanski B, Vaught LA, Reiser NL, et al. Low Levels of Neutralizing Antibodies After Natural Infection With Severe Acute Respiratory Syndrome Coronavirus 2 in a Community-Based Serological Study. Open Forum Infect Dis. 2022;9(3):ofac055.

11. Alfego D, Sullivan A, Poirier B, Williams J, Adcock D, Letovsky S. A population-based analysis of the longevity of SARS-CoV-2 antibody seropositivity in the United States. EClinicalMedicine. 2021;36:100902.

12. Decru B, Van Elslande J, Steels S, Van Pottelbergh G, Godderis L, Van Holm B, et al. IgG Anti-Spike Antibodies and Surrogate Neutralizing Antibody Levels Decline Faster 3 to 10 Months After BNT162b2 Vaccination Than After SARS-CoV-2 Infection in Healthcare Workers. Front Immunol. 2022;13:909910.

13. Yang Y, Yang M, Peng Y, Liang Y, Wei J, Xing L, et al. Longitudinal analysis of antibody dynamics in COVID-19 convalescents reveals neutralizing responses up to 16 months after infection. Nat Microbiol. 2022;7(3):423–33.

14. Wang Z, Muecksch F, Schaefer-Babajew D, Finkin S, Viant C, Gaebler C, et al. Naturally enhanced neutralizing breadth against SARS-CoV-2 one year after infection. Nature. 2021;595(7867):426–31.

15. Pérez-Alós L, Armenteros JJA, Madsen JR, Hansen CB, Jarlhelt I, Hamm SR, et al. Modeling of waning immunity after SARS-CoV-2 vaccination and influencing factors. Nat Commun. 2022;13(1):1614.

16. Addetia A, Crawford KHD, Dingens A, Zhu H, Roychoudhury P, Huang ML, et al. Neutralizing Antibodies Correlate with Protection from SARS-CoV-2 in Humans during a Fishery Vessel Outbreak with a High Attack Rate. J Clin Microbiol. 2020;58(11).

17. Khoury DS, Cromer D, Reynaldi A, Schlub TE, Wheatley AK, Juno JA, et al. Neutralizing antibody levels are highly predictive of immune protection from symptomatic SARS-CoV-2 infection. Nat Med. 2021;27(7):1205–11.

18. Lumley SF, O’Donnell D, Stoesser NE, Matthews PC, Howarth A, Hatch SB, et al. Antibody Status and Incidence of SARS-CoV-2 Infection in Health Care Workers. N Engl J Med. 2021;384(6):533–40.

19. Clarke KEN, Jones JM, Deng Y, Nycz E, Lee A, Iachan R, et al. Seroprevalence of Infection-Induced SARS-CoV-2 Antibodies - United States, September 2021-February 2022. MMWR Morb Mortal Wkly Rep. 2022;71(17):606–8.

20. Arkell P, Gusmao C, Sheridan SL, Tanesi MY, Gomes N, Oakley T, et al. Serological surveillance of healthcare workers to evaluate natural infection- and vaccine-derived immunity to SARS-CoV-2 during an outbreak in Dili, Timor-Leste. Int J Infect Dis. 2022;119:80–6.

21. Batchi-Bouyou AL, Djontu JC, Vouvoungui JC, Mfoutou Mapanguy CC, Lobaloba Ingoba L, Mougany JS, et al. Assessment of neutralizing antibody responses after natural SARS-CoV-2 infection and vaccination in congolese individuals. BMC Infect Dis. 2022;22(1):610.

22. Bonura F, Genovese D, Amodio E, Calamusa G, Sanfilippo GL, Cacioppo F, et al. Neutralizing Antibodies Response against SARS-CoV-2 Variants of Concern Elicited by Prior Infection or mRNA BNT162b2 Vaccination. Vaccines (Basel). 2022;10(6).

23. Altarawneh HN, Chemaitelly H, Ayoub HH, Tang P, Hasan MR, Yassine HM, et al. Effects of Previous Infection and Vaccination on Symptomatic Omicron Infections. N Engl J Med. 2022;387(1):21–34.

24. Gazit S, Shlezinger R, Perez G, Lotan R, Peretz A, Ben-Tov A, et al. SARS-CoV-2 Naturally Acquired Immunity vs. Vaccine-induced Immunity, Reinfections versus Breakthrough Infections: a Retrospective Cohort Study. Clin Infect Dis. 2022.

25. Lumley SF, Rodger G, Constantinides B, Sanderson N, Chau KK, Street TL, et al. An Observational Cohort Study on the Incidence of Severe Acute Respiratory Syndrome Coronavirus 2 (SARS-CoV-2) Infection and B.1.1.7 Variant Infection in Healthcare Workers by Antibody and Vaccination Status. Clin Infect Dis. 2022;74(7):1208–19.

26. Hall V, Foulkes S, Insalata F, Kirwan P, Saei A, Atti A, et al. Protection against SARS-CoV-2 after Covid-19 Vaccination and Previous Infection. N Engl J Med. 2022;386(13):1207–20.

27. Harvala H, Robb ML, Watkins N, Ijaz S, Dicks S, Patel M, et al. Convalescent plasma therapy for the treatment of patients with COVID-19: Assessment of methods Available for antibody detection and their correlation with neutralising antibody levels. Transfus Med. 2021;31(3):167–75.

28. Legros V, Denolly S, Vogrig M, Boson B, Siret E, Rigaill J, et al. A longitudinal study of SARS-CoV-2-infected patients reveals a high correlation between neutralizing antibodies and COVID-19 severity. Cell Mol Immunol. 2021;18(2):318–27.

29. Rijkers G, Murk JL, Wintermans B, van Looy B, van den Berge M, Veenemans J, et al. Differences in Antibody Kinetics and Functionality Between Severe and Mild Severe Acute Respiratory Syndrome Coronavirus 2 Infections. J Infect Dis. 2020;222(8):1265–9.

30. Scheiblauer H, Nübling CM, Wolf T, Khodamoradi Y, Bellinghausen C, Sonntagbauer M, et al. Antibody response to SARS-CoV-2 for more than one year - kinetics and persistence of detection are predominantly determined by avidity progression and test design. J Clin Virol. 2022;146:105052.

31. McCallum M, Czudnochowski N, Rosen LE, Zepeda SK, Bowen JE, Walls AC, et al. Structural basis of SARS-CoV-2 Omicron immune evasion and receptor engagement. Science. 2022;375(6583):864–8.

32. Maxmen A. Why call it BA.2.12.1? A guide to the tangled Omicron family. Nature. 2022;606(7914):446–7.

33. Yao L, Zhu KL, Jiang XL, Wang XJ, Zhan BD, Gao HX, et al. Omicron subvariants escape antibodies elicited by vaccination and BA.2.2 infection. Lancet Infect Dis. 2022;22(8):1116–7.

